# Is there evidence of inequality in the provision, level, and timing of SEND provision in English primary schools?

**DOI:** 10.1101/2025.08.27.25334554

**Authors:** Bianca L De Stavola, Vincent G. Nguyen, Kate Lewis, Andrea Aparicio Castro, Lorraine Dearden, Ruth Gilbert, the HOPE study team

## Abstract

Delays and inequalities in access to Special Educational Needs and Disability (SEND) provision in early childhood are likely to alter children’s social and education trajectories long-term. Effective policy solutions require robust evidence on the timing, duration, and contexts in which children with early indicators of need can access SEND provision.

We conducted a longitudinal study of 983,652 singleton children born in England in 2006-08 using ECHILD, a database containing linked hospital and state-funded school records. Nearly 16% of the study population had recorded SEND provision in Year 1 (age 5/6 years; 14.1% SEN Support, provided to children with less severe difficulties; 1.7% Education and Health Care Plan (EHCP), intended for those with more severe difficulties). SEN Support was more prevalent among males, those with pre-existing chronic health conditions and characterised by disadvantage and low Early Year Foundation Stage profile (EYFSP) scores. EHCP followed similar patterns but associations with disadvantage were weaker. Voluntary and academy sponsor-led schools were less likely to provide EHCPs, once known imbalances in pupil intake were accounted for.

Children entering state-funded education in nursery had higher prevalence of SEND provision throughout primary school than later entrants. EHCP increased gradually while SEN Support rose slowly, peaking at Year 2 (age 6/7 years) and then flattening. Children clustered into four trajectories of SEND provision from Year 1 to 6 labelled “*Never*”, “*Stable-High*”, “*Decreasing*” and “*Increasing”.* Those in the “*Never*” trajectory (77%) tended to live in income advantaged areas, had lower levels of chronic health conditions and higher EYFSP scores than peers. Children in the “*Increasing*” (i.e. delayed) trajectory were more likely to attend academy sponsor-led schools.

Results suggest that there are inequalities in type and timing of SEND provision that are associated with school governance.

## Introduction

In England, children are legally allowed to attend school from September after they turn four years old (Reception class) but must begin school in the term after they turn five [1, 2] with primary education, 95% of which is provided by the state sector [3], continuing from Year 1 (age 5/6) to Year 6 (age 10/11). For many children, however, education starts earlier, in nursery or other early years settings (if aged two or three).

Early years provision is largely (∼70%; [4]) delivered by private, voluntary, and independent providers, with state funding subsidising a set number of free childcare hours. Since 2010, most children aged three to four are entitled to 15–30 hours of free provision per week for 38 weeks annually [5], while access for two- to three-year-olds is more restricted. State funded nurseries primarily serve children with significant social or educational needs [6, 7]; they are commonly -but not always-integrated within local authority (LA) maintained primary schools (“Community schools”; see Appendix 1). In contrast, Academy and Foundation primary schools, which are also state funded but independent of LA control, are not obliged to provide free nursery places, nor special educational needs and disabilities (SEND) support [4, 8, 9, 10]. Thus, state funded, LA maintained, nurseries are the main setting for early years SEND support. Similarly, although all state funded (primary and secondary) schools must offer provision for children with SEND, those that are not LA maintained have been found to operate different threshold for SEND provision [11, 12]. Within this context, securing equitable and consistent access to SEND provision -fundamental to the Equality Act (2010) [13] and the SEND Code of Practice (2015) [14] - presents significant challenges.

Since SEND can impact a child’s learning through behavioural, cognitive, physical, or mental difficulties, provision in school takes different formats: SEN Support, consisting of additional attention given in class (e.g. by a support teacher) for children with less severe difficulties; and Education, Health and Care Plan (EHCP) intended to provide for children with more severe difficulties. SEND Support is funded through schools’ delegated budgets, while EHCPs are jointly funded by schools and LAs. Determining the appropriate level often involves multiple stakeholders—including health, social care, and education professionals—and is frequently perceived as inequitable [15, 16]. Parents’ and carers’ ability to navigate the system, particularly for EHCPs, further contributes to inconsistencies in access to provision [17, 18].

Understanding the factors that determine if, when, where, how, and for how long SEND support is provided is therefore essential. This paper aims to provide evidence to aid such understanding drawing on a birth cohort of children born 2006–2008 from the ECHILD database who attended Year 1 in state-funded schools in England between September 2012 and August 2014, and for whom we have proxy measures of need derived from health and social characteristics, as well as data on recorded SEND provision up to Year 6.

## Methods

### Data sources and Study Population

We created a birth cohort of children born between 1 September 2006 and 31 August 2008 using linked administrative data from the ECHILD (Education and Child Health Insights from Linked Data) database [19] as part of the HOPE Study (Health Outcomes of young People throughout Education Study; [20]). ECHILD includes hospital episode statistics admitted patient care (HES APC) data (see Appendix 1) linked to education data from the National Pupil Database (NPD) and to death registrations from the Office for National Statistics (ONS). HES APC holds data on births, diagnoses and procedures recorded by NHS funded hospitals in England for children born in England since 1997 while NPD contains termly information on school registration, absences, and exclusions for children attending state funded schools in England from the academic year 2001/02, while attainment is available yearly [21]. Information on the children’s birth characteristics are available from the birth records held in HES and on socio-demographic characteristics from both HES and NPD. Data on SEND provision recorded in state funded nurseries and state funded primary education are also available in NPD. Of note, such records do not necessarily indicate that SEND provision was received, nor is there any information about the precise elements of support received (hence the use hereafter of *‘*recorded SEND provision’).

We constructed the study population by including all live singleton children in ECHILD who were born between 1 September 2006 and 31 August 2008 and who entered Year 1 (the first year of compulsory education) in a state primary school during the 2012/2013 or 2013/2014 academic year. This birth cohort was selected to ensure that follow-up did not extend beyond 2019, thereby avoiding potential biases arising from the COVID-19 pandemic, which substantially disrupted school attendance, SEND provision, and data quality. We excluded children with inconsistent, incomplete, or invalid dates of birth across databases. The remaining records were linked to the Early Years Census (EYC) [23], which contains information on attendance and recorded SEND provision in private or voluntary nurseries, as well as in maintained nurseries not attached to a school.

### Data

The variables derived from HES, NPD and EYC are described in Box 1. The information on SEND provision concerns each year from Nursery to Reception (when applicable), and then from Year 1 to Year 6. It was derived by appending data from EYC to data to the NPD spring census (the one deemed to be more complete).

##### Description of the variables used and of their sources

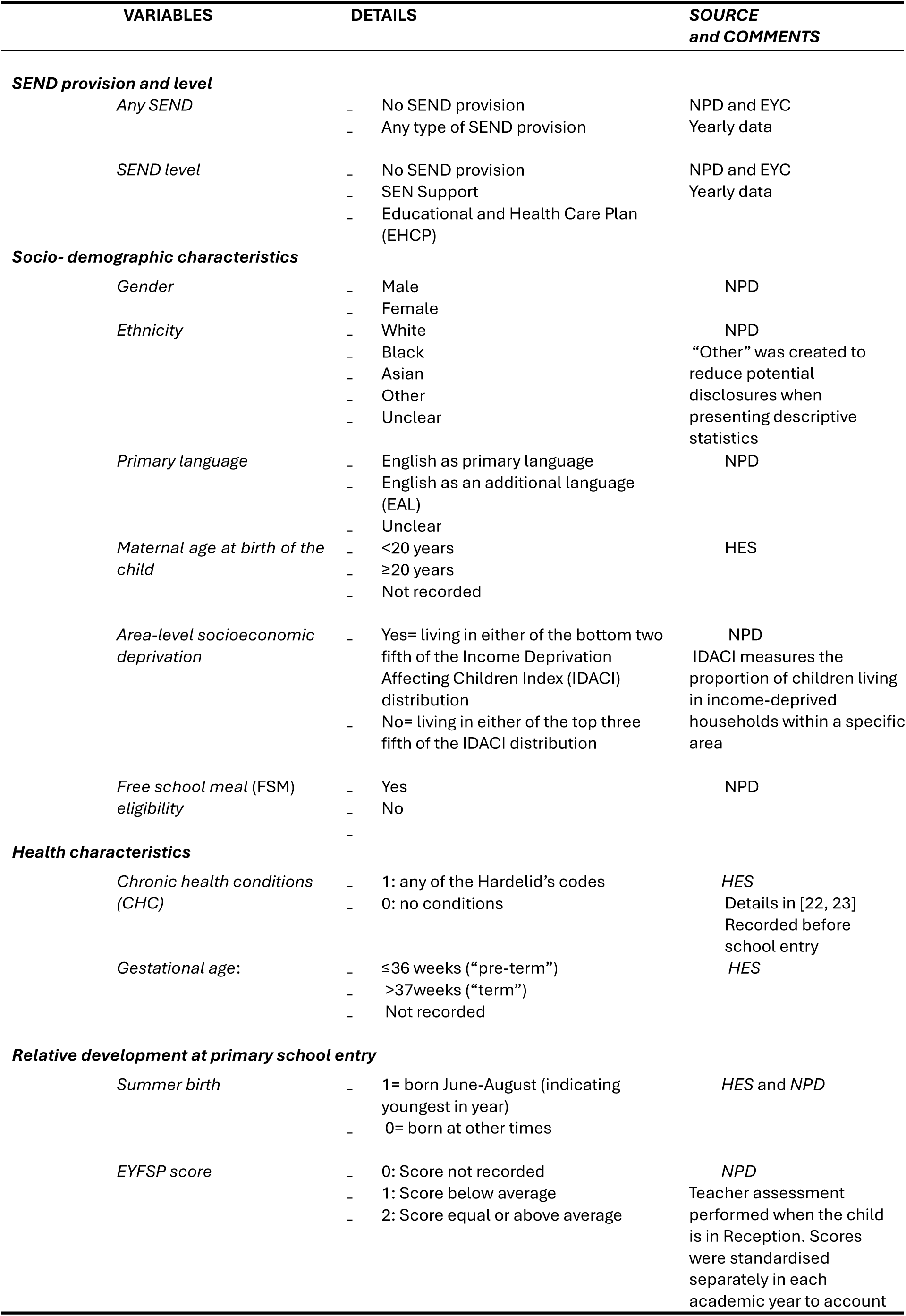
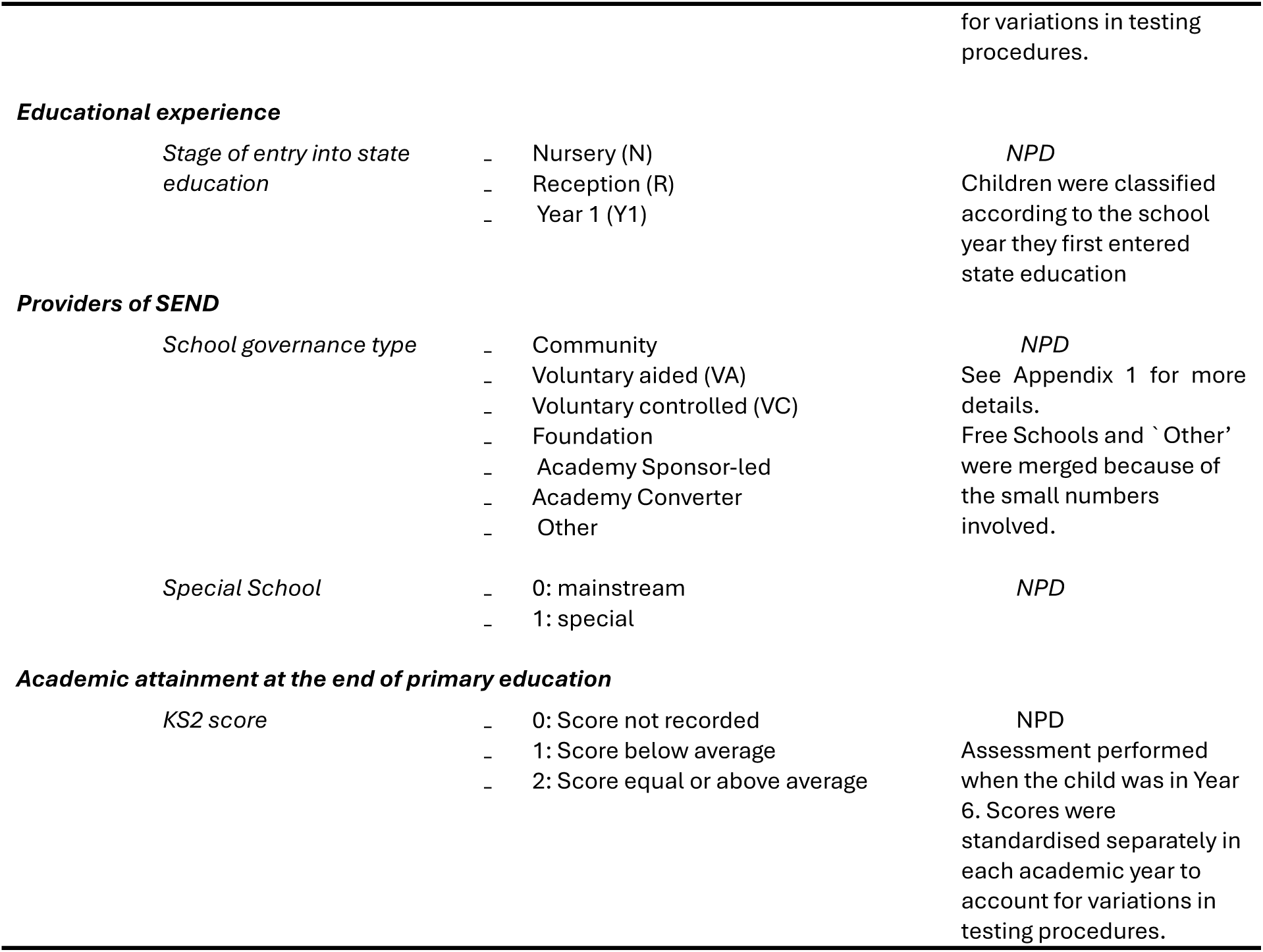

BOX 1 – Description of the variables used and of their sources
| VARIABLES | DETAILS | SOURCE<br>and COMMENTS |
| --- | --- | --- |
| **SEND provision and level** |  |  |
| *Any SEND* | - No SEND provision<br>- Any type of SEND provision | NPD and EYC<br>Yearly data |
| *SEND level* | - No SEND provision<br>- SEN Support<br>- Educational and Health Care Plan (EHCP) | NPD and EYC<br>Yearly data |
| **Socio- demographic characteristics** |  |  |
| *Gender* | - Male<br>- Female | NPD |
| *Ethnicity* | - White<br>- Black<br>- Asian<br>- Other<br>- Unclear | NPD<br>“Other” was created to reduce potential disclosures when presenting descriptive statistics |
| *Primary language* | - English as primary language<br>- English as an additional language (EAL)<br>- Unclear | NPD |
| *Maternal age at birth of the child* | - <20 years<br>- ≥20 years<br>- Not recorded | HES |
| *Area-level socioeconomic deprivation* | - Yes= living in either of the bottom two fifth of the Income Deprivation Affecting Children Index (IDACI) distribution<br>- No= living in either of the top three fifth of the IDACI distribution | NPD<br>IDACI measures the proportion of children living in income-deprived households within a specific area |
| *Free school meal (FSM) eligibility* | - Yes<br>- No<br>- | NPD |
| **Health characteristics** |  |  |
| *Chronic health conditions (CHC)* | - 1: any of the Hardelid’s codes<br>- 0: no conditions | HES<br>Details in [22, 23]<br>Recorded before school entry |
| *Gestational age:* | - ≤36 weeks (“pre-term”)<br>- >37weeks (“term”)<br>- Not recorded | HES |
| **Relative development at primary school entry** |  |  |
| *Summer birth* | - 1= born June-August (indicating youngest in year)<br>- 0= born at other times | HES and NPD |
| *EYFSP score* | - 0: Score not recorded<br>- 1: Score below average<br>- 2: Score equal or above average | NPD<br>Teacher assessment performed when the child is in Reception. Scores were standardised separately in each academic year to account |
|  |  | for variations in testing procedures. |
| **Educational experience** |  |  |
| Stage of entry into state education | - Nursery (N)<br>- Reception (R)<br>- Year 1 (Y1) | *NPD*<br>Children were classified according to the school year they first entered state education |
| **Providers of SEND** |  |  |
| School governance type | - Community<br>- Voluntary aided (VA)<br>- Voluntary controlled (VC)<br>- Foundation<br>- Academy Sponsor-led<br>- Academy Converter<br>- Other | *NPD*<br>See Appendix 1 for more details.<br>Free Schools and 'Other' were merged because of the small numbers involved. |
| Special School | - 0: mainstream<br>- 1: special | *NPD* |
| **Academic attainment at the end of primary education** |  |  |
| KS2 score | - 0: Score not recorded<br>- 1: Score below average<br>- 2: Score equal or above average | *NPD*<br>Assessment performed when the child was in Year 6. Scores were standardised separately in each academic year to account for variations in testing procedures. |

### Statistical Methods

We first described the study population and then the proportions of recorded SEND provision in Year 1 (by category of SEND provision) in terms of the socio-demographic and health characteristics listed above, and of the indicators of relative development by Year 1 and stage of entry into state education. We then considered whether SEND provision varied by school governance type, once differences in pupils’ intake were accounted for. To study SEND provision over time we used longitudinal data on SEND provision from Nursery to Year 6 to compare groups of children with different stages at entry into state education and then modelled the data from Year 1 to Year 6 to identify, and then compare, the most typical trajectories of provision.

More specifically, these analyses consisted of the following steps:

#### 1. Description of the study population

We calculated the percentage distribution among the study participants of their socio-demographic and health characteristics first, and then also of their relative development at start of compulsory school and of stage at entry.

#### 2. SEND provision in Year 1

The percentage distribution of any SEND provision and, separately, of SEND Support and EHCP, recorded at the start of the first full year of compulsory education (i.e. Year 1) were calculated for each of the socio-demographic and health characteristics, relative development and stage at entry, and reported graphically. We also reported the percentage distribution of SEND provision by providers of SEND.

To assess whether provision differed according to type of school governance, we estimated the average predicted probability of recorded SEND in Year 1 by school governance type. These are the probabilities that would occur if all children had attended each of the school type [24]. We achieved this by:

a. Fitting a logistic regression model for the outcome “Any SEND” that included the socio-demographic and health characteristics, relative development and stage at entry variables described above;
b. Estimating the predicted probabilities specific to each school governance for each child in the study population;
c. Averaging these predicted probabilities over the study population, thus achieving probabilities that are “*equalised*” over the distribution of the covariates used in (a).

We repeated these steps for categorical SEND (with categories of “No SEND”, “SEN Support” and “EHCP”) fitting a multinomial regression model in (a), instead of logistic regression. We compared the equalised probabilities with the observed frequencies of pupils across school governance type. Results are reported graphically.

#### 3. Cross-sectional trajectories of SEND provision during primary education

To investigate temporal changes in SEND provision, we first calculated the percentage of pupils with any SEND provision (and, separately, with SEN Support and EHCP) for each school year from the start of their educational experience up to the end of primary education. These percentages were calculated separately by groups defined by stage of entry into state education to highlight whether differentials in average trajectories were present. These plots exploit the linked information on SEND provision during state funded hours in private or voluntary nurseries recorded in the EYC. For this reason, lines that are specific to a stage of entry can start earlier than the entry time into the state system.

#### 4. Individual trajectories of SEND provision during primary education

We used the individually linked longitudinal information on whether children had recorded any SEND provision and modelled these data using latent class longitudinal models [25] to examine whether histories of SEND provision clustered into separate groups from Year 1 to 6. We examined overall SEND provision because the numbers with EHCP were too few.

The number of groups (latent classes) was selected comparing results of alternative models with an increasing number of classes; the model which provided the smallest value of the Bayesian Information Criterion (BIC) when the size of the smallest class was >5% was selected. Each child was assigned to their most likely class according to their predicted probabilities of belonging to each class. The results are shown as plots of the estimated latent trajectory classes. In a final step, we related the most likely class of each child to their socio-demographic, health, relative development characteristics and stage at entry. We also re-examined whether there was evidence of differentials according to school governance.

### Missing values

Missing values affected some of the variables sourced from birth records: gestational age was missing for 34.8% of records, while maternal age was missing only for 3.6%. When fitting regression models to derive equalised probabilities we used the missing value indicator method to retain the records with incomplete data, aware that some confounding bias will remain when this method is used with observational data, but adopting it to deal with the larger bias that would occur if dropping the records [26].

## Results

### 1. Study Population

A total of 1,014,145 live singleton births recorded in NHS hospitals in England between 1 September 2006 and 31 August 2008 were linked to NPD. To avoid incorrect linkages, we excluded children whose dates of birth were inconsistent or incomplete across databases or were inadmissible, leaving 1,010,556 children, and then excluded further 191 whose school year was inconsistent with the reported calendar year. We also excluded children whose first record in NPD referred to Year 2 or later or who did not have any records in Year 1. This led to a study population of 983,652 children (Figure 1), most of whom entered the state school system in Reception (age 4/5; 566,283; 57.6%), with the second largest group consisting of those who entered in Nursery (age 2-3; 413,340; 42.0%). Very few were first enrolled in Year 1 (age 5/6; 4,029; 0.4%).

**Figure 1.**
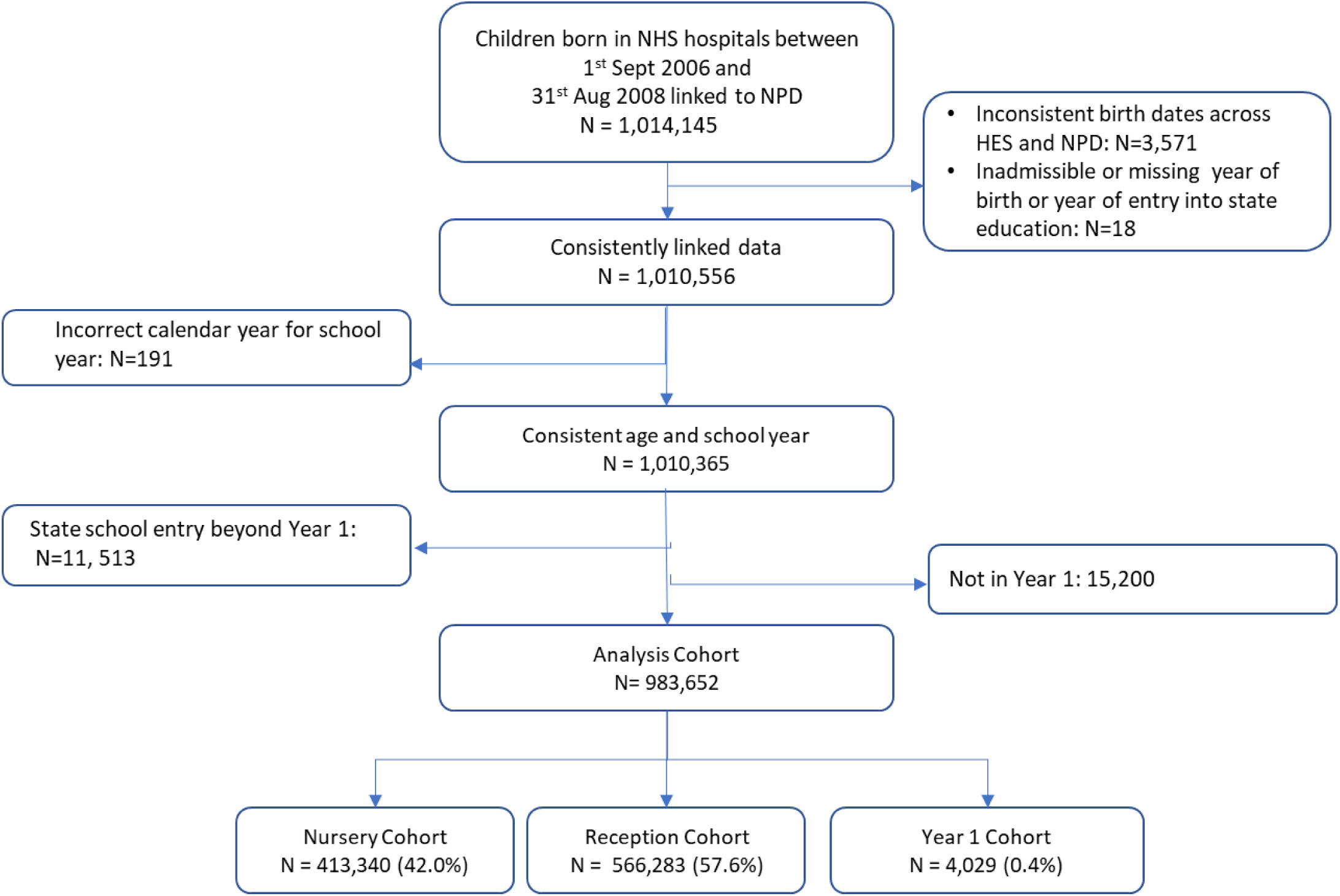
Derivation of the HOPE study cohort of children born in 2006-08

### 2. SEND provision in Year 1

SEND provision (of any type) during Year 1 was recorded for nearly 16% of all children (N= 155,064, 15.8%), with EHCP recorded for 1.7% and SEN Support for 14.1% (Table 1). Provision percentages varied by socio-demographic and health characteristics (Table 1; Figure 2). Boys had twice the recorded SEND provision of girls (21.0% vs 10.2%) as did those with pre-existing chronic health conditions (CHCs) compared to those without (26.6% vs 13.4%). Children born prematurely had around 60% more SEND provision recorded than those born at term (24.0% vs 15.2%). Similar patterns of higher prevalence of recorded SEND provision were seen for the indicators of socio-economic disadvantage: those living in more deprived areas, those whose mother was <20-year-old at their birth, and those eligible for free school meal (FSM) had greater – up to twice - the percentage of SEND provision of those without these features (Table 1, Figure 2(a)). The prevalence of SEND provision in Year 1 was highest among Black children and those with English as an additional language (EAL).

**Figure 2.**
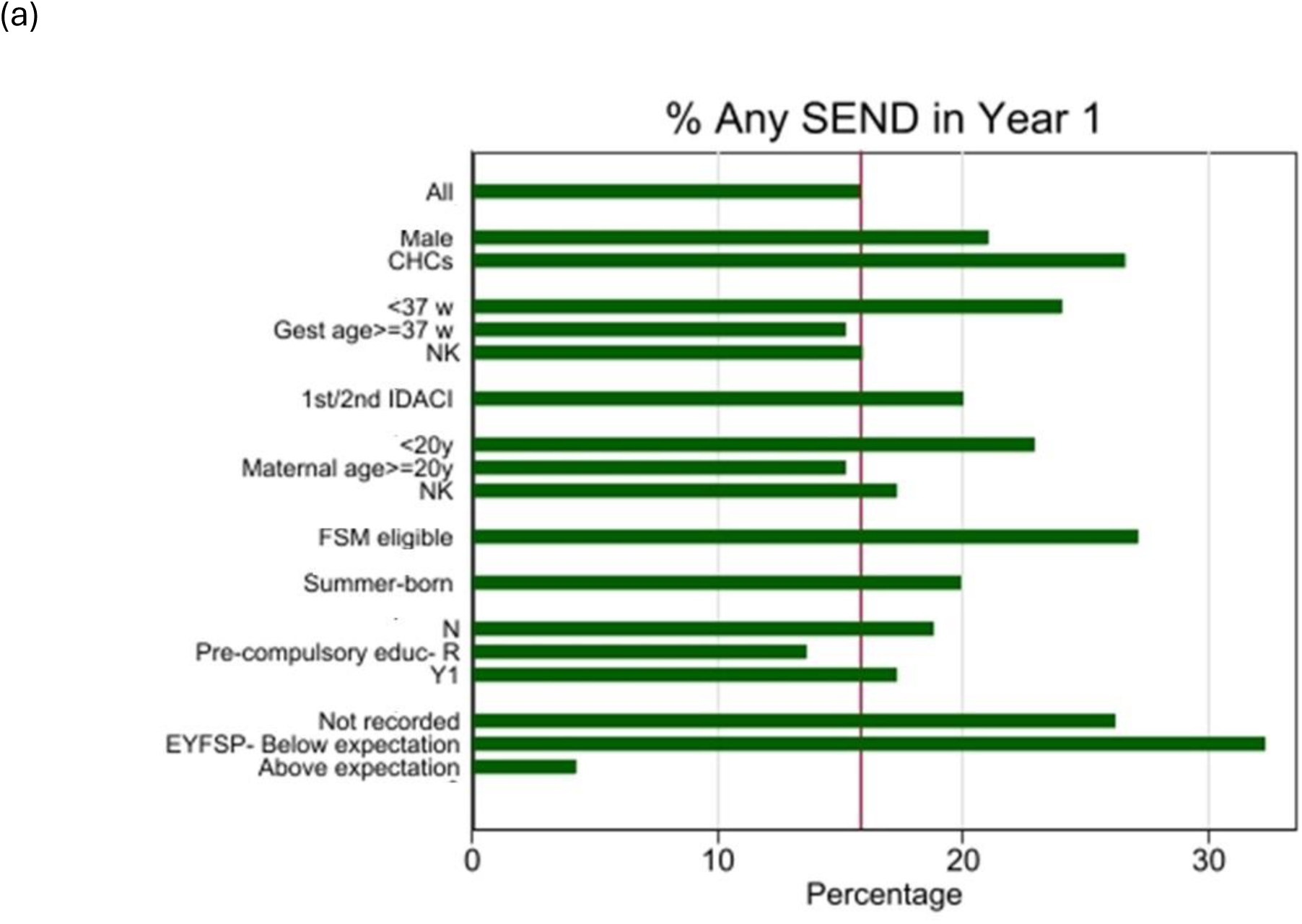
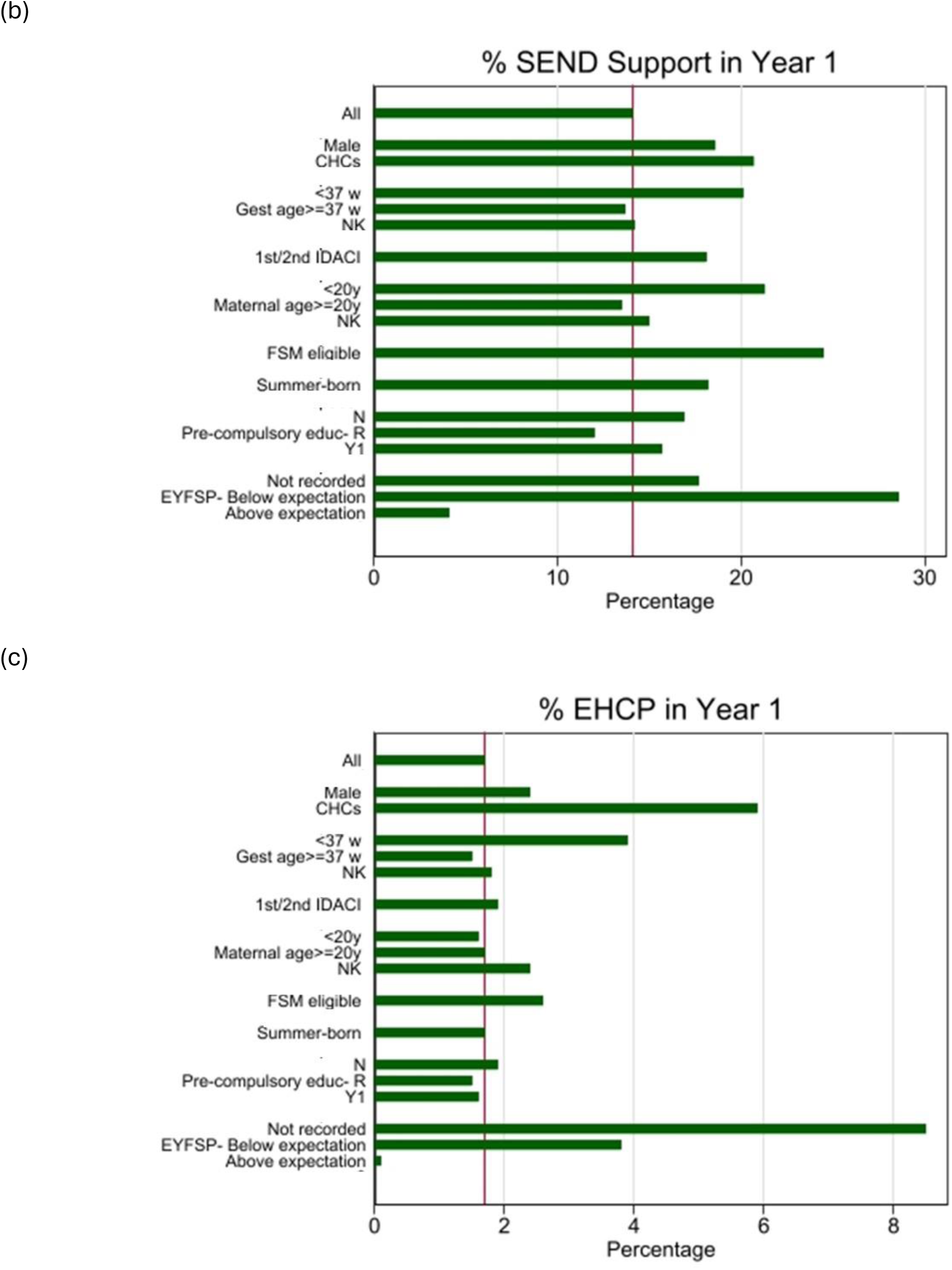
Percentage of SEND provision by socio-demographic and relative development indicators: (a) Any SEND; (b) SEN Support; (c) EHCP; HOPE cohort of children born in 2006-08

**Table 1.** Pupils’ characteristics and their recorded SEND provision in Year 1; HOPE cohort of children born in 2006-08, N= 983,652.

|  | Total |  | Any SEND |  | SEN Support |  | EHCP |  |
| --- | --- | --- | --- | --- | --- | --- | --- | --- |
|  | N | % | N | % | N | % | N | % |
| **All** | 983,652 | **100.0** | 155,064 | **15.8** | 138,556 | **14.1** | 16,508 | **1.7** |
| **Gender** |  |  |  |  |  |  |  |  |
| Male | 506,070 | **51.5** | 106,208 | **21.0** | 94,226 | **18.6** | 11,982 | **2.4** |
| Female | 477,582 | **48.5** | 48,856 | **10.2** | 44,330 | **9.3** | 4,526 | **1.0** |
| **CHCs** |  |  |  |  |  |  |  |  |
| Any recorded | 178,162 | **18.1** | 47,424 | **26.6** | 36,878 | **20.7** | 10,546 | **5.9** |
| Not recorded | 805,490 | **81.9** | 107,640 | **13.4** | 101,678 | **12.6** | 5,962 | **0.7** |
| **Gestational age** |  |  |  |  |  |  |  |  |
| <37 weeks | 37,584 | **3.8** | 9,030 | **24.0** | 7,558 | **20.1** | 1,472 | **3.9** |
| ≥37weeks | 604,286 | **61.4** | 91,546 | **15.2** | 82,543 | **13.7** | 9,003 | **1.5** |
| NK | 341,782 | **34.8** | 54,488 | **15.9** | 48,455 | **14.2** | 6,033 | **1.8** |
| **IDACI** |  |  |  |  |  |  |  |  |
| 1<sup>st</sup>/2<sup>nd</sup> quintile | 464,519 | **47.2** | 92,825 | **20.0** | 83,945 | **18.1** | 8,880 | **1.9** |
| 3<sup>rd</sup> /5<sup>th</sup> quintile | 519,133 | **52.8** | 62,239 | **12.0** | 54,611 | **10.5** | 7,628 | **1.5** |
| **Maternal Age at delivery** |  |  |  |  |  |  |  |  |
| <20 years | 64,889 | **6.6** | 14,833 | **22.9** | 13,809 | **21.3** | 1,024 | **1.6** |
| ≥20 years | 883,293 | **89.8** | 134,082 | **15.2** | 119,432 | **13.5** | 14,650 | **1.7** |
| NK | 35,470 | **3.6** | 6,149 | **17.3** | 5,315 | **15.0** | 834 | **2.4** |
| **FSM eligibility** |  |  |  |  |  |  |  |  |
| yes | 187,570 | **19.1** | 50,814 | **27.1** | 45,906 | **24.5** | 4,908 | **2.6** |
| no | 796,082 | **80.9** | 104,250 | **13.1** | 92,650 | **11.6** | 11,600 | **1.5** |
| **Ethic group** |  |  |  |  |  |  |  |  |
| White | 757,461 | **77.0** | 116,876 | **15.4** | 104,996 | **13.9** | 11,880 | **1.6** |
| Black | 52,084 | **5.3** | 10,045 | **19.3** | 8,762 | **16.8** | 1,283 | **2.5** |
| Asian | 97,049 | **9.9** | 15,792 | **16.3** | 13,861 | **14.3** | 1,931 | **2.0** |
| Other | 75,120 | **7.6** | 12,077 | **16.1** | 10,705 | **14.3** | 1,372 | **1.8** |
| Unclear | 1,938 | **0.2** | 274 | **14.1** | 232 | **12.0** | 42 | **2.2** |
| **Main Language** |  |  |  |  |  |  |  |  |
| English | 822,695 | **83.6** | 127,482 | **15.5** | 114,131 | **13.9** | 13,351 | **1.6** |
| EAL | 158,910 | **16.2** | 27,257 | **17.2** | 24,141 | **15.2** | 3,116 | **2.0** |
| Unclear | 2,047 | **0.2** | 325 | **15.9** | 284 | **13.9** | 41 | **2.0** |
| **Summer born** |  |  |  |  |  |  |  |  |
| Yes | 256,742 | **26.1** | 51,036 | **19.9** | 46,765 | **18.2** | 4,271 | **1.7** |
| No | 726,910 | **73.9** | 104,028 | **14.3** | 91,791 | **12.6** | 12,237 | **1.7** |
| **EYFSP score** |  |  |  |  |  |  |  |  |
| Not recorded | 11,265 | **1.2** | 2,951 | **26.2** | 1,992 | **17.7** | 959 | **8.5** |
| Below average | 396,578 | **40.3** | 128,096 | **32.3** | 113,206 | **28.6** | 14,890 | **3.8** |
| Above average | 575,809 | **58.5** | 24,017 | **4.2** | 23,358 | **4.1** | 659 | **0.1** |
| **Stage at start of state education** |  |  |  |  |  |  |  |  |
| Nursery | 413,342 | **42.0** | 77,533 | **18.8** | 69,765 | **16.9** | 7,758 | **1.9** |
| Reception | 566,281 | **57.6** | 76,843 | **13.6** | 68,158 | **12.0** | 8,685 | **1.5** |
| Year 1 | 4,029 | **0.4** | 698 | **17.3** | 633 | **15.7** | 65 | **1.6** |

*CHC: chronic health conditions; EAL: English as an alternative language; EYFSP: Early years foundation stage profile; IDACI: Income Deprivation Affecting Children Index*

Children born in summer had about 40% greater frequency of recorded SEND provision in Year 1 than those born at other times, and those with missing or below average EYFSP scores had up to nearly 8 times the provision of the others. For children who entered state education in Nursery, SEND provision was about 40% higher than for those entering in Reception (Table 1; similar raised percentages were seen for the very small number of children who entered in Year 1). This reflects the relative characteristics of these children: those entering in Reception are the least likely group to have pre-existing CHCs or to be characterised by disadvantaged conditions (Suppl Table 1).

In contrast, the distributions in recorded EHCP in Year 1, which involves only 1.7% of children, differ with respect to several factors (Table 1; Figure 2(c)). Specifically, those with mothers younger than 20 years did not have higher EHCP percentages than the rest; very few children with above average EYFSP scores had an EHCP and there were no substantial differences in EHCP frequencies according to season of birth. Entering state education in Year 1 did not increase the frequency of EHCP, in contrast to SEN Support, but this may reflect the time needed to obtain access to the former. Interestingly, children with pre-existing CHCs, who are a fifth (18.1%) of all children in the study population, accounted for two thirds (63.9%) of those with an EHCP, while less than a third (26.6%) of those with SEN Support did so (30.6% overall, i.e. for any SEND).

In Year 1, which for this cohort was attended in 2012/13 or 2013/14, most (58.0%) children attended Community schools, i.e. schools controlled by the LA (Table 2). Voluntary Controlled schools and Foundation schools are also controlled by the LA but have more autonomy: they were attended in Year 1 by, respectively, 9.2% and 5.7% of pupils. Voluntary Aided and Academy schools, although also state funded, are not controlled by the LA. They served respectively 17.1% and 9.6% of pupils. The two types of Academies are very distinctive, however: most Academy sponsor-led are Community schools that were underperforming and were forced to change status, while Academy Converter are schools that were high-performers and that decided to gain independence from their LA. The remaining schools (in the “Other” category) encompasses only 0.3% of pupils. Provision of any SEND, and specifically EHCP, in Year 1 varied greatly across these schools: Voluntary Aided and Voluntary Controlled schools had the lowest percentages of all children overall (13.1% and 12.5% vs. the average of 15.8%; Table 2), as well as lowest percentages of EHCP provision (0.9% and 1% vs. the overall average of 1.7%). The highest percentages for any type of SEND are found for Academy Sponsor-led and Community schools (20.5% and 16.9%); for EHCP provision the highest percentages are those for Community and Foundation schools (2.0% and 1.7%), as well as for schools in the “Other” category (4.0%). This category however included the largest proportion of special school pupils across all school types (5.0% vs 0.6% overall).

**Table 2.** Frequency of SEND providers by recorded SEND provision in Year 1; HOPE cohort of children born in 2006-08, N= 983,652.

|  | TOTAL |  | Any SEN |  | SEN Support |  | EHCP |  |
| --- | --- | --- | --- | --- | --- | --- | --- | --- |
|  | N | % | N | % | N | % | N | % |
| **All** | 983,652 | **100.0** | 155,064 | **15.8** | 138,556 | **14.1** | 16,508 | **1.7** |
| **School governance** |  |  |  |  |  |  |  |  |
| Community | 570,409 | **58.0** | 96,342 | **16.9** | 84,887 | **14.9** | 11,455 | **2.0** |
| Voluntary Aided | 168,629 | **17.1** | 22,149 | **13.1** | 20,705 | **12.3** | 1,444 | **0.9** |
| Voluntary Controlled | 90,841 | **9.2** | 11,357 | **12.5** | 10,494 | **11.6** | 863 | **1.0** |
| Foundation | 56,158 | **5.7** | 9,301 | **16.6** | 8,180 | **14.6** | 1,121 | **2.0** |
| Academy Sponsor-led | 25,936 | **2.6** | 5,325 | **20.5** | 4,975 | **19.2** | 350 | **1.4** |
| Academy Converter | 69,144 | **7.0** | 10,116 | **14.6** | 8,942 | **12.9** | 1,174 | **1.7** |
| Other | 2,535 | **0.3** | 474 | **18.7** | 373 | **14.7** | 101 | **4.0** |
| **Special school** |  |  |  |  |  |  |  |  |
| yes | 5,900 | **0.6** | 5,900 | **100.0** | 112 | **1.9** | 5,788 | **98.1** |
| no | 977,752 | **99.4** | 149,164 | **15.3** | 138,444 | **14.2** | 10,720 | **1.1** |

These variations in SEND provision reflect the pupil characteristics of the schools, as highlighted in Figure 3. Community and, especially, Academy Sponsor-led schools were attended in Year 1 more frequently by pupils characterised by socio-economic disadvantage (lived in the most deprived areas, were born to younger mothers, and were FSM eligible). Their pupils were also less likely to be of White ethnicity, or to have English as primary language, or to have above average EYFSP scores.

**Figure 3.**
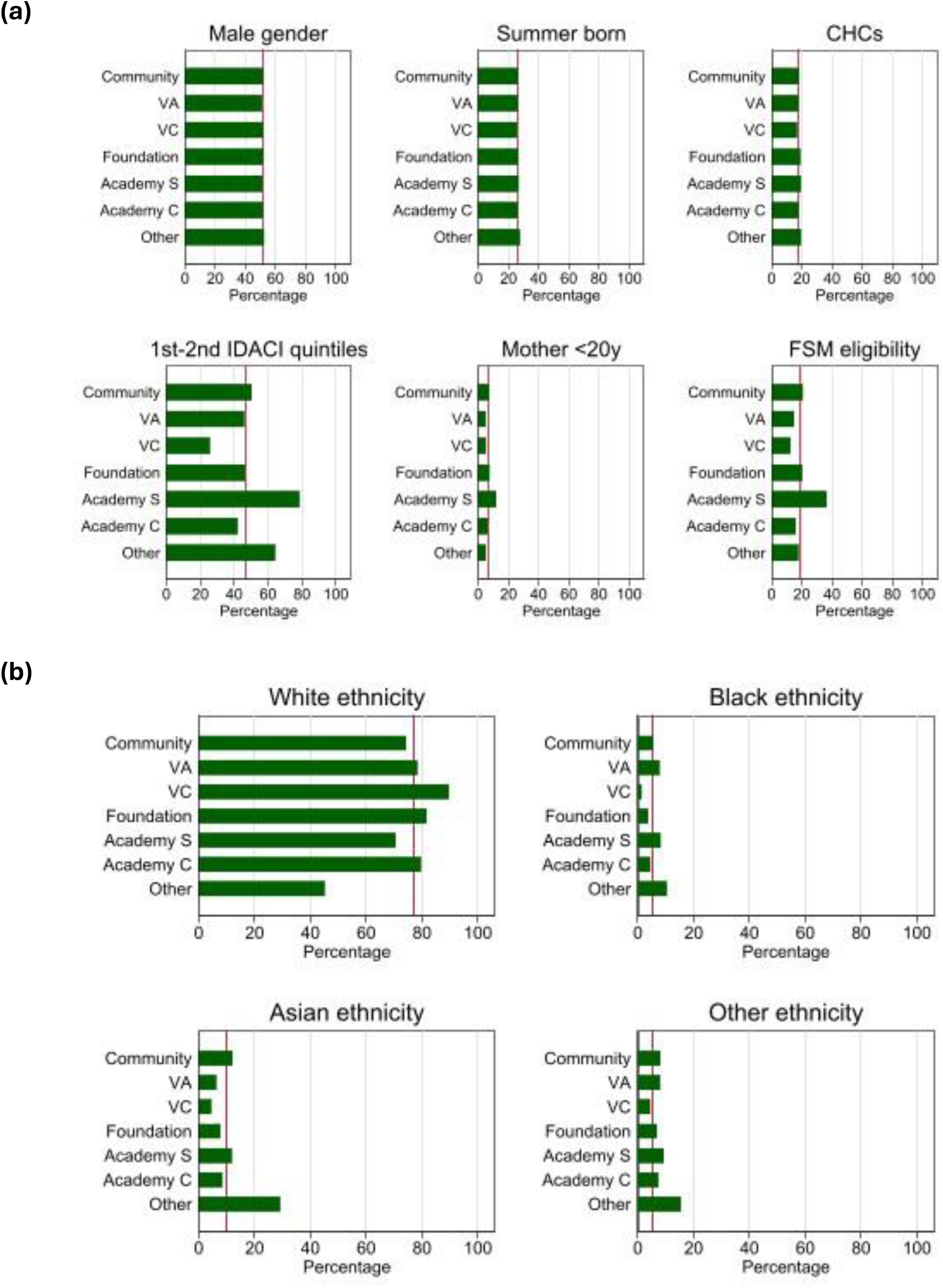
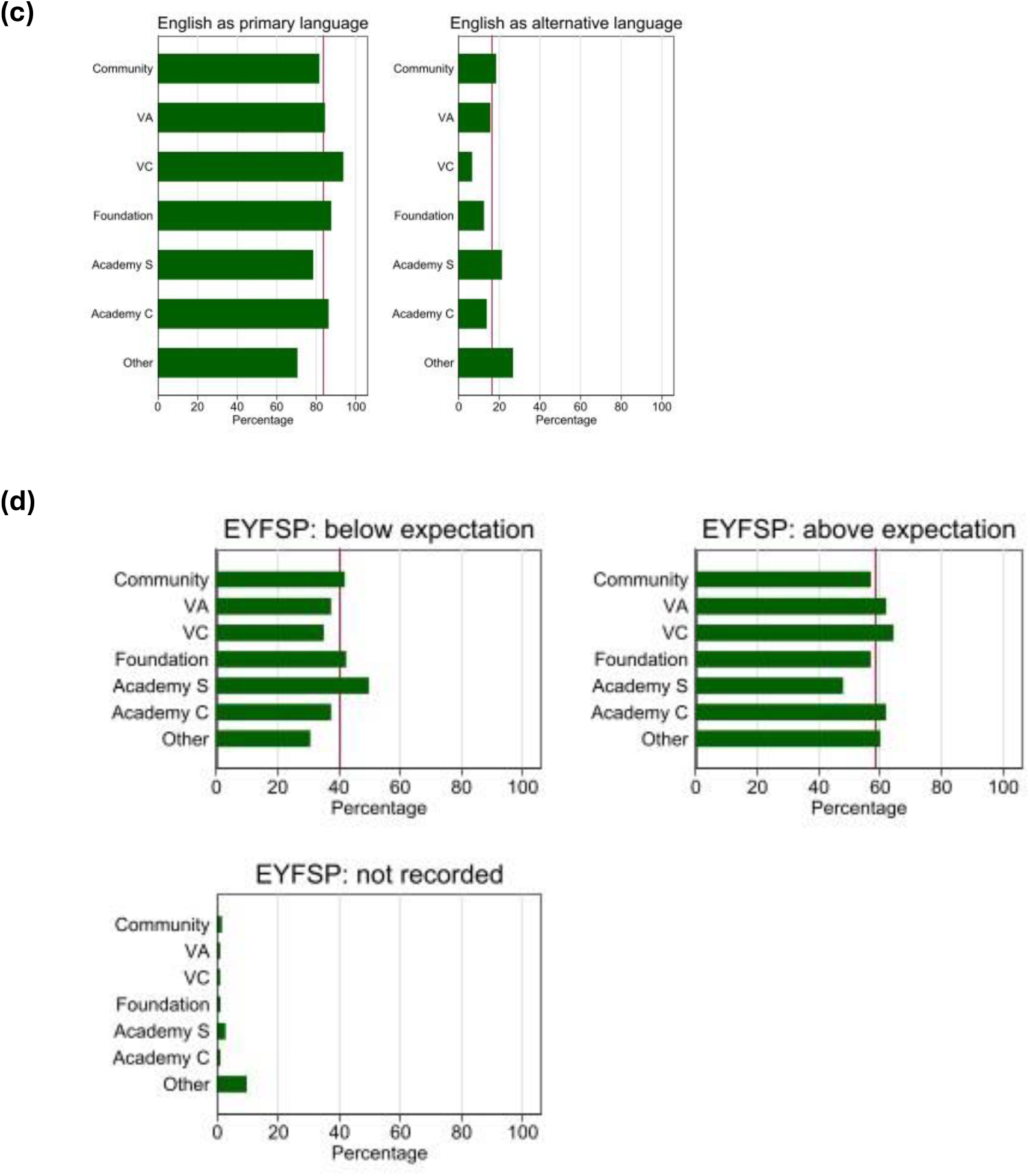
Pupils’ characteristics by school governance type: (a) socio-demographic; (b) ethnicity; (c) primary language; (d) EYFSP

To examine whether there observed differentials in SEND provision by school governance type were indicative not just of their differences in children’s intake but also of systematic inequalities, we estimated what the probability of being assigned to SEND (SEN Support and specifically EHCP) would be, if their pupils’ characteristics were equalised. Figure 4(a) shows both observed and equalised probabilities of SEN Support by school governance type. The equalised proportions all converge towards the overall mean. However, when this exercise was repeated just for EHCP provision, the equalised probabilities were still substantially smaller than the average specifically for Voluntary Aided schools, Voluntary Controlled and - surprisingly given their overall SEND frequencies - Academy Sponsor-led schools (Figure 4(b)).

**Figure 4.**
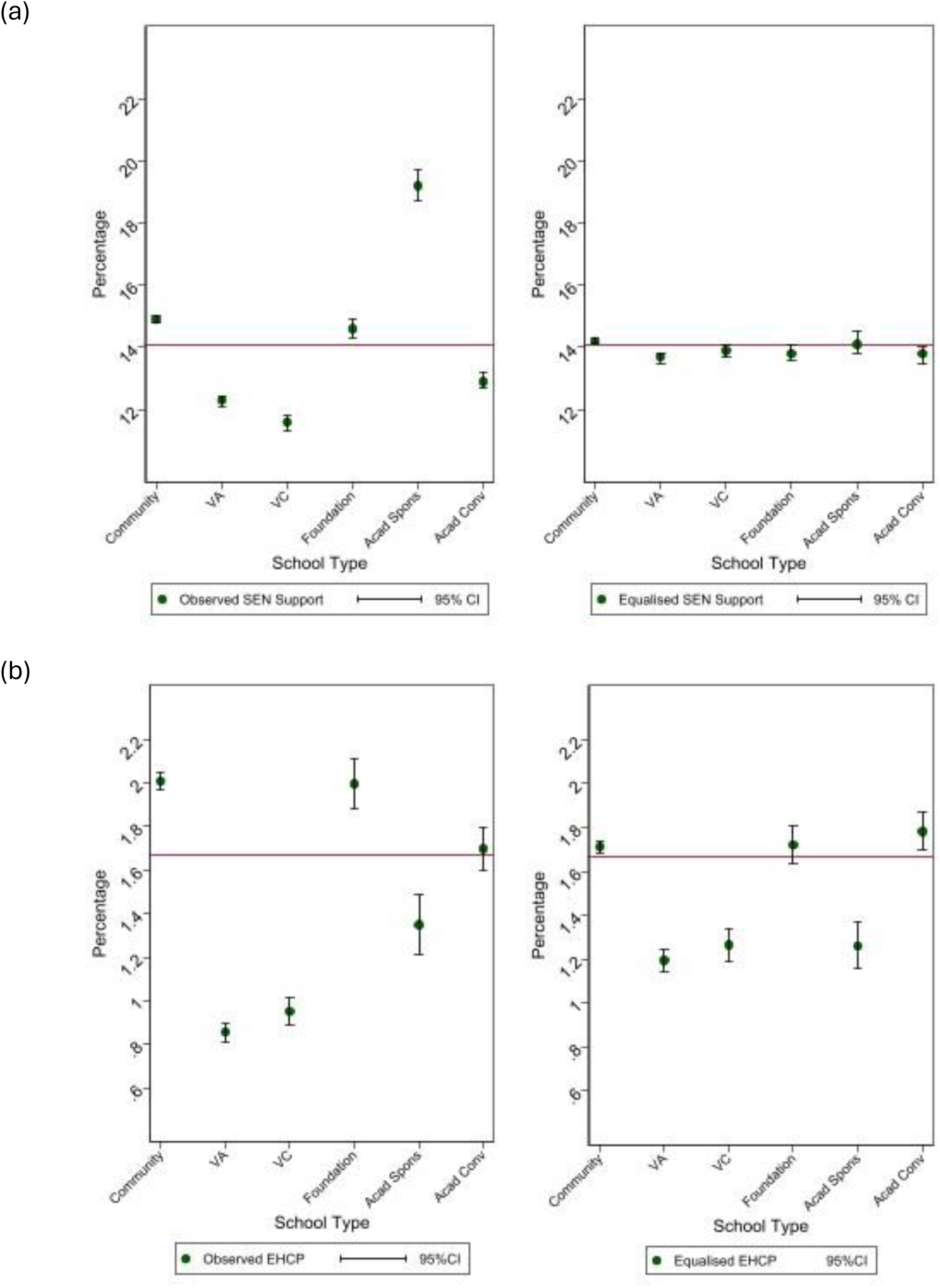
Observed and equalised percentage of SEND provision by school governance type: (a) SEN Support; (b) EHCP

### 3. Cross-sectional trajectories of SEND provision during primary education

SEND provision varied over the school years, as shown in Figure 5, where the percentages of SEN Support and of EHCP provision are reported for each academic year from Nursery to the end of primary education, Year 6. The percentages were calculated separately by stage of entry into state education and highlight substantial differentials in SEN Support (black lines): those who started in Nursery had the highest percentages from the start, while those who started in Reception had the lowest, in line with the earlier observation about their characteristics. In both cases, percentages increase from the early years up to Year 2, then both lines flatten (with some decrease and then increase, Figure 5). For the very small proportion of children who entered the state education system in Year 1 (0.4% of the total), SEN Support was much higher than for the rest from the time of their entry in Year 1 (there were no records of earlier SEND in EYC for them), increasing then very sharply up to Year 2 before slightly decreasing.

**Figure 5.**
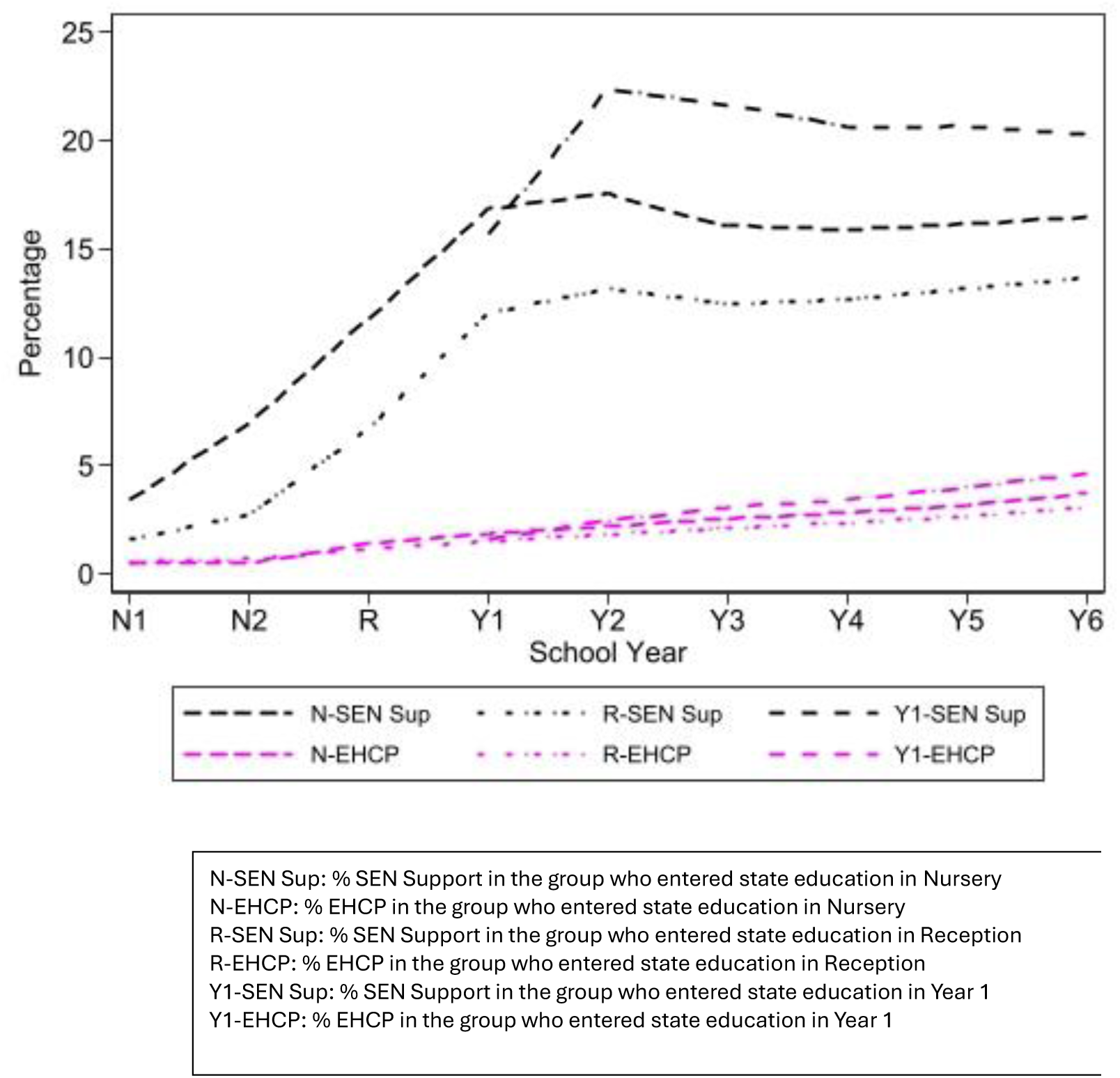
Percentage SEN Support and EHCP pupils from nursery by stage at entry into state education

Different trajectories are seen for recorded EHCP (pink lines). Although they are close to zero in the Nursery years, they increase steadily over time, with the children who entered state education in Reception having the lowest and those entering in Year 1 the highest proportions.

In Year 6 the proportion of children with recorded SEN Support was respectively 16.4% for those who entered in Nursery, 13.7% for those who entered in Reception and 20.3% for those who entered in Year 1. The respective percentages for EHCP were: 3.7%, 3.1% and 4.7%. Thus, from Year 1 to Year 6 EHCP provision doubles while SEN Support hardly increases (cf. Table 1).

### 4. Individual trajectories of SEND provision during primary education

The percentages reported in Figure 5 were derived separately for each school year. To study instead trajectories of SEND provision as experienced by each child, we fitted a set of multilevel models to the longitudinal data ranging from Year 1 to Year 6, the end of primary education. Initial results provided an estimate of the average SEND history for these children. However, the heterogeneity in observed SEND provision over time led us to fit models that allow for different average trajectories, i.e. latent class latent trajectory models [25]. We compared results across models that allowed different numbers of trajectories, including the simpler version with a single average trajectory, and found that the best fitting model (see Suppl Table 2) had four typical trajectories of SEND provision characterised by different shapes and frequencies (Figure 6). The first (Class 1, C1), which we labelled “*Never*”, included 77.1% of the children in the study population and is characterised by very low probabilities of SEND provision; the second class (C2), labelled “*Stable-high*”, involved 11.8% of the children and is characterised by consistently high SEND provision from Year 1; the third class (C3, “I*ncreasing*”) and the fourth class (C4, “*Decreasing*”) involved respectively 5.7% and 5.6% of children, with the first showing a gradual increase in SEND provision from Year 2, and the second displaying an initial high probability of SEND followed by a notable decrease from Year 2.

**Figure 6.**
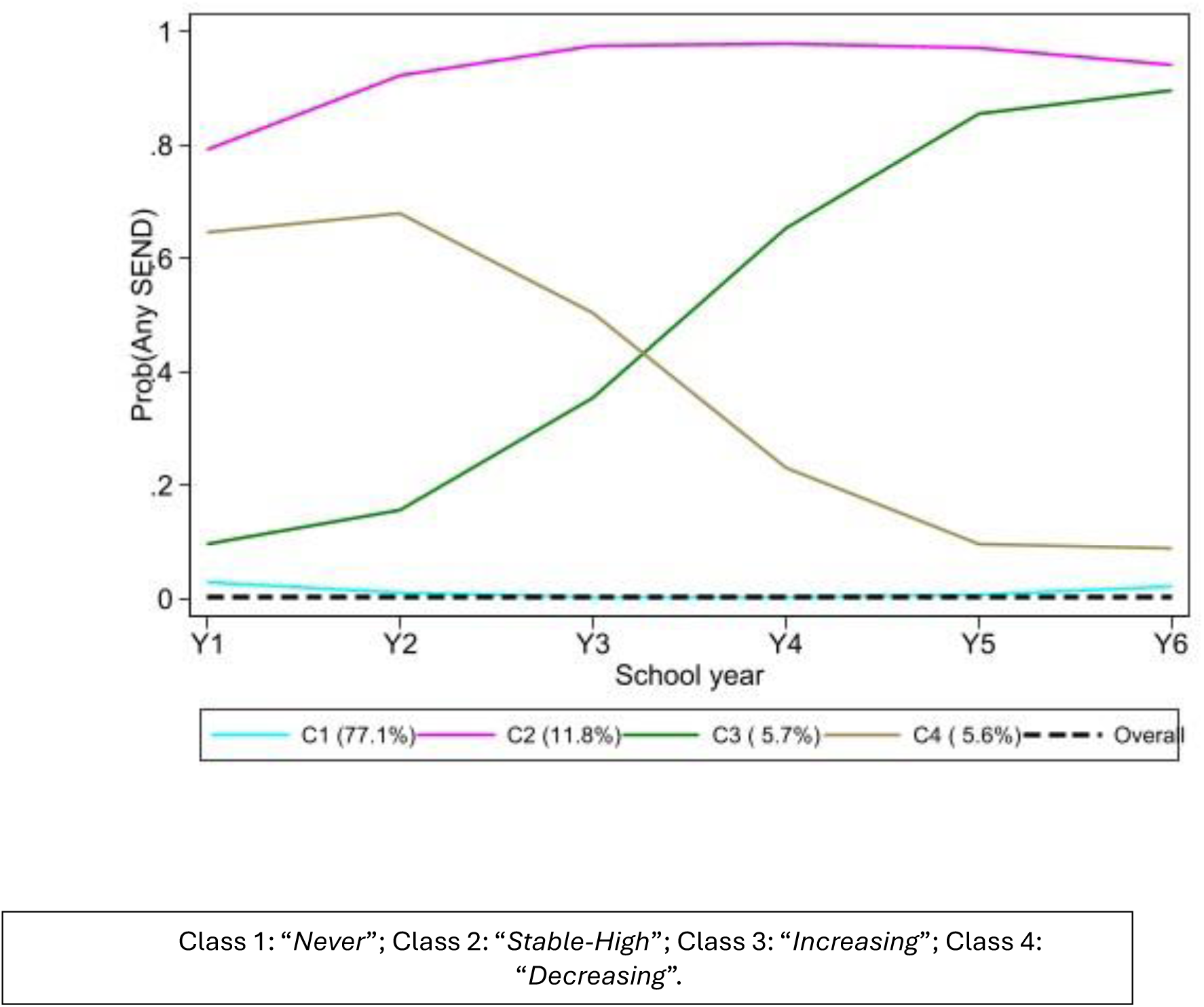
Estimated trajectories of any SEND provision according to the four latent classes identified by the best fitting latent class growth model

Children following these four trajectories are likely to differ in terms of their needs, both at entry into and during their primary education. We do not have access to measures of need for SEND but can examine the socio-demographic, health and developmental indicators at the beginning of the trajectories, i.e. in Year 1, separately by trajectory class (Table 3). Children assigned to C1 (*Never*) were more likely to be female (53% vs the overall average of 48.5%-see Table 1), to live in more affluent areas (55.7% vs 52.8%), and not to be FSM eligible (84.7% vs 80.9%). Children assigned to C2 (*Stable-high*) were more likely to be male (70.4% vs 51.5%), to have pre-existing CHCs (34.0% vs 18.1%) and to have been born pre-term (6.2% vs 3.8%). They also lived in more disadvantaged areas (58.4% vs 47.2%) and were more likely to be FSM eligible (34.4% vs 19.1%). Children assigned to C3 (*Increasing*) were again more likely to be male (60.6% vs 51.5%) to have CHCs (22.0% vs 18.1%), to live in more disadvantaged areas (52.4% vs 47.2%) and to be FSM eligible (28.4% vs 19.1%) but, for each of these features, percentages were not as large as those characterizing C2 children. However, children in C3 were more likely to be White (81.9% vs 77.0%) and to have English as their primary language (89.0% vs 83.6%) than children in C2. Those assigned to C4 (*Decreasing*) were children with the highest frequencies of living in deprived areas (59.0% vs 47.2%), of being of Black or Asian ethnicity (17.3% vs 15.2%), and of having EAL (19.3% vs 16.2%). They also had the highest proportion of summer births (34.0% vs 26.1%).

**Table 3.**
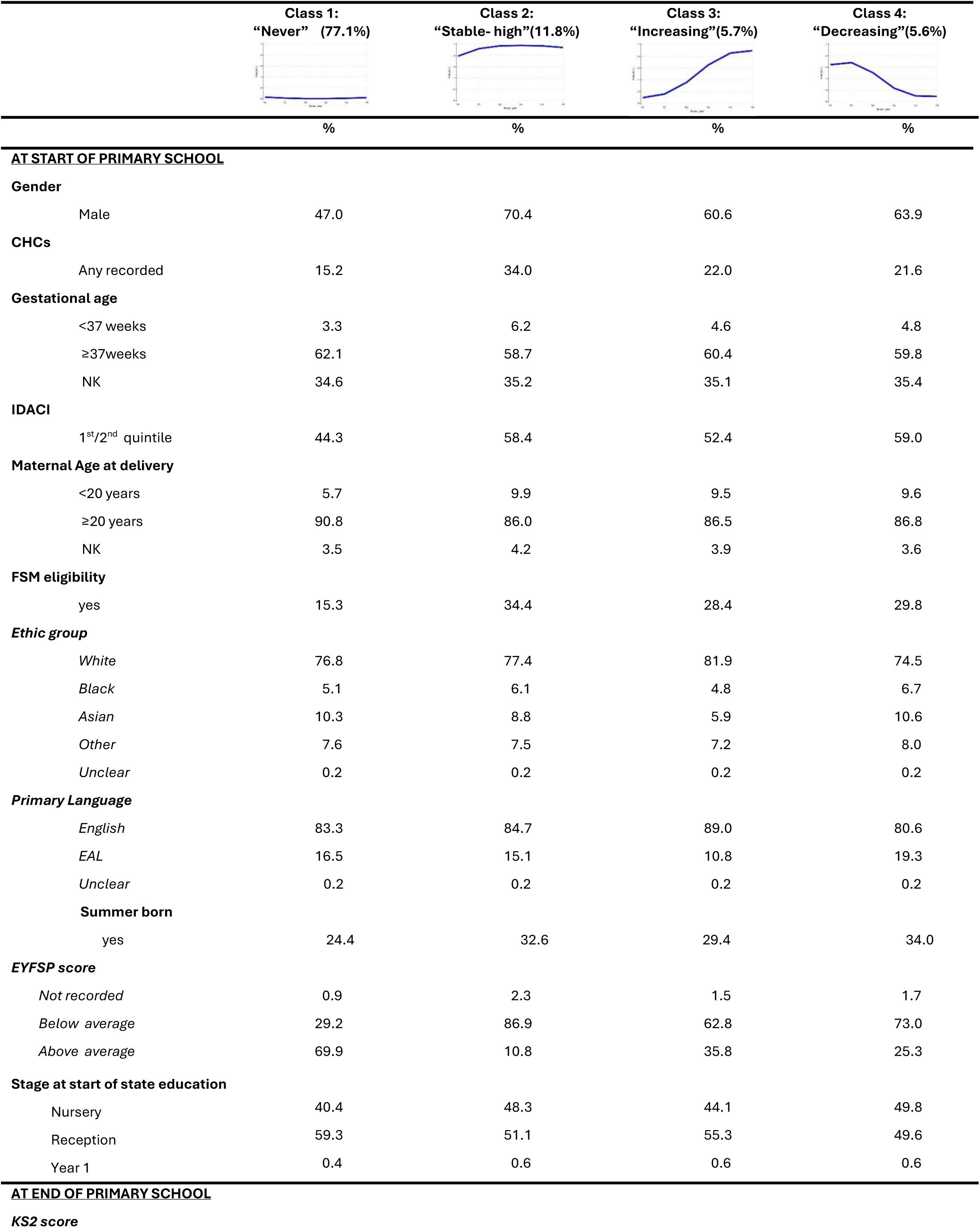
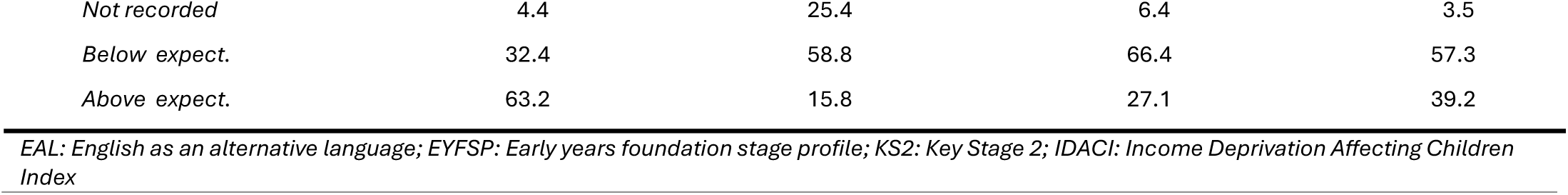
Pupils’ characteristics at entry and end of primary state education by SEND trajectory class; HOPE cohort of children born in 2006-08, N= 983,652.

|  | Class 1:<br>“Never” (77.1%) | Class 2:<br>“Stable- high”(11.8%) | Class 3:<br>“Increasing”(5.7%) | Class 4:<br>“Decreasing”(5.6%) |
| --- | --- | --- | --- | --- |
|  | % | % | % | % |
| **AT START OF PRIMARY SCHOOL** |  |  |  |  |
| **Gender** |  |  |  |  |
| Male | 47.0 | 70.4 | 60.6 | 63.9 |
| **CHCs** |  |  |  |  |
| Any recorded | 15.2 | 34.0 | 22.0 | 21.6 |
| **Gestational age** |  |  |  |  |
| <37 weeks | 3.3 | 6.2 | 4.6 | 4.8 |
| ≥37weeks | 62.1 | 58.7 | 60.4 | 59.8 |
| NK | 34.6 | 35.2 | 35.1 | 35.4 |
| **IDACI** |  |  |  |  |
| 1<sup>st</sup>/2<sup>nd</sup> quintile | 44.3 | 58.4 | 52.4 | 59.0 |
| **Maternal Age at delivery** |  |  |  |  |
| <20 years | 5.7 | 9.9 | 9.5 | 9.6 |
| ≥20 years | 90.8 | 86.0 | 86.5 | 86.8 |
| NK | 3.5 | 4.2 | 3.9 | 3.6 |
| **FSM eligibility** |  |  |  |  |
| yes | 15.3 | 34.4 | 28.4 | 29.8 |
| **Ethnic group** |  |  |  |  |
| White | 76.8 | 77.4 | 81.9 | 74.5 |
| Black | 5.1 | 6.1 | 4.8 | 6.7 |
| Asian | 10.3 | 8.8 | 5.9 | 10.6 |
| Other | 7.6 | 7.5 | 7.2 | 8.0 |
| Unclear | 0.2 | 0.2 | 0.2 | 0.2 |
| **Primary Language** |  |  |  |  |
| English | 83.3 | 84.7 | 89.0 | 80.6 |
| EAL | 16.5 | 15.1 | 10.8 | 19.3 |
| Unclear | 0.2 | 0.2 | 0.2 | 0.2 |
| **Summer born** |  |  |  |  |
| yes | 24.4 | 32.6 | 29.4 | 34.0 |
| **EYFSP score** |  |  |  |  |
| Not recorded | 0.9 | 2.3 | 1.5 | 1.7 |
| Below average | 29.2 | 86.9 | 62.8 | 73.0 |
| Above average | 69.9 | 10.8 | 35.8 | 25.3 |
| **Stage at start of state education** |  |  |  |  |
| Nursery | 40.4 | 48.3 | 44.1 | 49.8 |
| Reception | 59.3 | 51.1 | 55.3 | 49.6 |
| Year 1 | 0.4 | 0.6 | 0.6 | 0.6 |
| **AT END OF PRIMARY SCHOOL** |  |  |  |  |
| **KS2 score** |  |  |  |  |
| *Not recorded* | 4.4 | 25.4 | 6.4 | 3.5 |
| *Below expect.* | 32.4 | 58.8 | 66.4 | 57.3 |
| *Above expect.* | 63.2 | 15.8 | 27.1 | 39.2 |

*EAL: English as an alternative language; EYFSP: Early years foundation stage profile; KS2: Key Stage 2; IDACI: Income Deprivation Affecting Children Index*

Unsurprisingly, children in C1 had the highest proportion of above average EYFSP scores (69.9% vs 58.5%; Table 3) and those in C2 had the lowest proportion (10.8%; they are also the only group with children attending special schools). Conversely, the EYFSP scores for children in C3 and C4 are quite similar (62.8% and 73%) while their trajectories are so different. The high frequency of summer-born children in C4 would be consistent with the shape of their trajectory, if a large component of them had caught-up with their peers by Year 2 and therefore were in decreasing need for SEND provision. However, there is no clear explanation for the delayed SEND provision experienced by C3 children, given their similarly poor EYFSP performance to those in C4. C3 is largely composed of children with White ethnicity whose primary language is English; they also were more likely to attend Academy Sponsor-led schools (Figure 7) and less likely to have entered state-education in Nursery than those in C4 (44.1% vs 49.8%; Table 3). Opportunities for their needs to be recognised early were therefore reduced, with delays in recognition and support likely to explain the differentials in academic performance at the end of primary education (Table 3). While the percentages of above average scores increased from EYFSP to KS2 for children in C4 (from 25.3% to 39.2%), they did not for those in C3 (going from 35.8% to 27.1%). The percentage of children with no recorded scores also increased more for C3 than C4 children.

**Figure 7.**
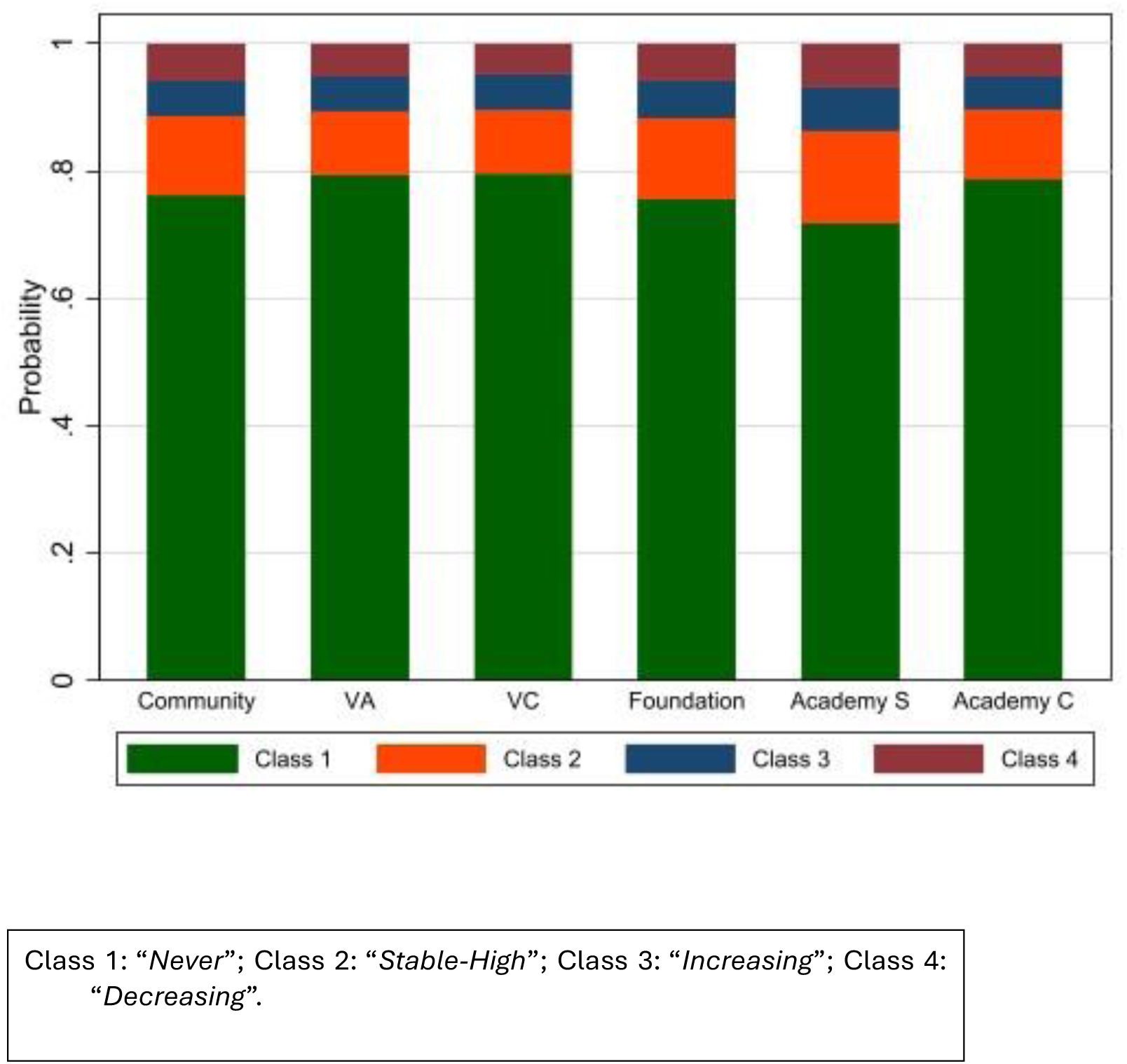
Observed distribution of trajectory classes by school governance type

## Discussion

### Overview of findings

Nearly 16% of the study population had recorded SEND provision in Year 1: 14.1% with SEN Support and 1.7% with an EHCP. The percentage of pupils with an EHCP doubled to 3.4% by the end of primary education but increased only moderately for SEN Support (to 14.9%). SEN Support was more prevalent among males, children with pre-existing CHCs and with indicators of social disadvantage (including having young mothers) or for pupils with low or missing EYFSP scores. EHCP provision followed similar patterns but had higher prevalences of children with CHCs or those born prematurely, while there were no differences between children of younger and older mothers. The associations with gender [27, 28, 29], disadvantage [30, 12, 31] and ethnicity [32, 33, 34] have been reported before. However, our highlighting that only one third of pupils with recorded SEND provision (and 64% for EHCP) had identified CHCs is novel.

The prevalence of SEN Support and EHCP provision in Year 1 varied across school governance types. For SEN Support the observed school differences disappeared once measured pupil characteristics were accounted for. However, differences persisted for EHCP provision, with lower equalised probabilities of provision found for children attending Voluntary and Academy Sponsor-led schools (state-funded schools outside of LA control). These results are partly in agreement with previous findings [12, 35, 36] but differ in flagging Voluntary schools, and in the finding that, despite offering SEN Support to large percentages of their pupils, Academy Sponsor-led schools had lower equalised probabilities of EHCP. In contrast Academy Converter schools had average equalised probabilities, reflecting the differences in pupils’ intake of the two types of Academy schools we found, as also discussed by Eyles (2018) [35, 37].

When studying SEND provision over time, four typical trajectories of SEND provision emerged: “*Never*”, “*Stable-High*”, “*Increasing*”, and “*Decreasing*”. Children assigned to the first two classes were characterised by opposite features: those assigned to the *Never* class were more often females and were characterised by more affluent conditions and higher EYFSP scores; those assigned to *Stable-High* were more often males and were characterised by disadvantaged conditions and very poor EYFSP scores. Children assigned to the *Increasing* and *Decreasing classes* were instead closer in terms of socio-economic disadvantage and EYFSP scores but differed in terms of ethnicity. Children assigned to the *Increasing* class were more likely to be of White ethnicity and to have English as their primary language; they were also more likely to attend Academy Sponsor-led schools.

The prevalence of summer births among the pupils in the *Decreasing* group could explain the downward trajectory, whereby earlier differences in need might reduce with time [38]. The group also included the largest percentage of children affected by conditions with the Hardelid’s respiratory code (such as cleft lip and/or palate), for whom needs for educational support are expected to fade [39]. The finding that White disadvantaged children attending certain schools are over-represented among those in the *Increasing* group, i.e. those for whom needs for SEND provision were recognised (or manifested) later, is however novel.

### Study design and limitations

Our analyses were based on a cohort of nearly 1 million children born in 2006-2008 who attended the first year of compulsory education in 2012/2013 or 2013/14 and for whom we have linked health and education data.

The 2006-08 birth period enabled full primary school follow-up, whilst avoiding the COVID-19 pandemic and providing follow-up data to the end of primary education for all children. This choice had however the disadvantage of focusing on a cohort that could have had a different experience of SEND provision from later cohorts because they attended Year 1 before the reforms of 2014 [35]. Generalizability to later cohorts therefore may be limited, although SEND provision during primary school has not changed greatly since 2013/14 (Suppl Figure 1), unlike what is seen for secondary schools [35]. The relative frequencies of school governance types, however, have changed, given the fast expansion of academies [35] which was also experienced by our cohort of children during their primary schooling (Suppl Figure 2). Therefore, our results, based on the schools attended in Year 1, may be underestimates of inequalities in SEND provision, given the increased prevalence of academy schools.

The most important limitation of our results, however, lies in the extent and quality of the information sourced from administrative databases and used in these analyses to proxy both the factors leading to SEND and SEND provision itself. Health indicators were derived from maternity and hospital diagnoses and were affected by missingness (for gestational age) and incompleteness (especially for illnesses that do not require hospitalization). Access to birth records, and hospitalization in infancy and childhood, did however add information on birth and acquired conditions that might affect need for SEND, and that were not available in similar studies based on linked NPD and census data only. As regards the main indicators of disadvantage, IDACI quintiles and FSM eligibility, although the former is area-based and can only be a proxy of a child’s conditions and the latter suffers from underreporting especially in younger pupils [31, 40], together they have been shown to capture underlying disadvantage [41]. Notably, the available information on SEND provision, sourced from NPD, refers only to recorded provision, not to whether SEND was delivered, nor its modality. Therefore, although the data allowed some, incomplete, control of underlying SEND, our reported comparisons of provision by school governance type concern recorded provision only.

### What is novel

This study refines and extends the evidence on SEND provision in primary education in England by summarising comprehensive data from pre-compulsory education through the end of primary school, disaggregated by stage of entry into state education.

Consistent with earlier research, we confirm associations between SEND provision and being male, experiencing poor health, or living in disadvantage [27, 28, 29, 30, 31, 33, 34]. In contrast to Hutchinson et al. [12, 36] who found evidence that children with EAL receive lower levels of provision, we report crude associations in the opposite direction; their findings however were based on modelled marginal probabilities derived from multilevel analyses that adjusted for multiple covariates. Given the complexities of structural and social factors that shape SEND provision, covariate adjustment may lead to spurious results if it includes variables that lie on the causal pathway from EAL to SEND (e.g. school absences), thus affecting the interpretation of the reported association.

We used multivariate models to estimate “equalised” probabilities of SEND provision (SEN Support and EHCP) by school governance type, where probabilities accounted for imbalances in pupils’ intake with the purpose if aiding fairness when comparing providers. Using longitudinal data, as recommended by Harland et al. [42], we identified four classes of common trajectories of SEND provision. These patterns align with qualitative findings [43, 44] that had previously not been quantified or systematically characterised.

Taken together, our results highlight inequalities in whether, when, how, and for how long SEND provision is experienced.

### Research and policy implications

The findings presented here underscore the need for more detailed data on SEND provision—specifically, refined information on whether, how, and when support is delivered, and on the factors most effective in identifying children in need. This study did not evaluate the extent to which SEND interventions benefit recipients. In earlier work, we explored this question within selected groups of children [45, 46], but found that available data, despite their richness, were insufficient to address causal questions. Such questions would require designs such as staggered randomised trials, for example comparing earlier provision with phased introduction. In the meantime, descriptive analyses such as those undertaken here can serve as tools for monitoring inequalities and delays in provision, of value to policymakers.

Finer data on the steps leading to the identification of SEND, on whether SEND is actually provided, and beyond that, on how SEND is provided, would allow robust insights into how education and health services collaborate in identifying children requiring support, and the quality and quantity of that support. Further data linkage on SEND provision between primary and secondary education would enhance the recent work by Hutchinson et al (2025) [36].

## Conclusions

We found evidence of inequalities in recorded EHCPs, as well as in the timing and duration of SEND provision, associated with stage of entry into state education and school governance type. These findings have important implications for policymakers, particularly in demonstrating how administrative data can be harnessed to monitor the delivery and equity of SEND provision.

## Abbreviations

EHCP: education and health care plan
EYC: early years census
EYFSP: early years foundation stage profile
ECHILD: Education and Child Health Insights from Linked Data
HES: hospital episodes statistics
HOPE: Health Outcomes of young People throughout Education
IDACI: Income Deprivation Affecting Children Index
KS2: key stage 2
LA: local authority
NPD: National Pupil Database
SEND: special education need and disabilities
SRS: secure research service

## Acknowledgements

Members of the HOPE study team include: Ruth Gilbert (PI), Katie Harron, Bianca L De Stavola, Lorraine Dearden, Tamsin Ford (senior work package leads), Kate Lewis, Vincent Nguyen, Ania Zylbersztejn, Jennifer Saxton, Jacob Matthews, William Farr (leading roles in the 4 work packages), Ayana Cant, Laura Gimeno, Isaac Winterburn,, Andrea Aparicio Castro, Julia Shumway, Lucy Karwatowska (contributing researchers), Matthew Lilliman (programme manager), Kate Boddy (public engagement coordinator), Stuart Logan, Jugnoo Rahi, Kristine Black-Hawkins, Johnny Downs (co-investigators).

We are grateful to the HOPE study Advisory Group: Chris Bonell (chair, professor of health and sociology, London School of Hygiene and Tropical Medicine), Kate Evans-Jones, Julia Ogden (public members), Jo Hutchison (Director of SEND, Education Policy Institute), Karen Horridge (consultant community paediatrician, Sunderland).

We gratefully acknowledge all children and families whose de-identified data were used in this research. We thank Ruth Blackburn, Milagros Ruiz, Matthew Jay, Antony Stone and Farzan Ramzan for ECHILD Database support.

The ECHILD Database uses data from the Department for Education (DfE). The DfE does not accept responsibility for any inferences or conclusions derived by the authors. This work contains statistical data from ONS which is Crown Copyright. The use of the ONS statistical data in this work does not imply the endorsement of the ONS in relation to the interpretation or analysis of the statistical data. This work uses research datasets which may not exactly reproduce National Statistics aggregates. Evidence from this research contributes to the NIHR Children and Families Policy Research Unit but was not commissioned by the NIHR Policy Research Programme.

## Ethics statement

Permissions to use linked, de-identified data from HES and NPD were granted by NHS England (DARS-NIC-381972-Q5F0V-v0.5) and the Department for Education (DR200604.02B). Ethical approval for the ECHILD project was granted by the National Research Ethics Service (17/LO/1494), NHS Health Research Authority Research Ethics Committee (20/EE/0180 and 21/SW/0159). Separate ethical approval was not required to use de-identified data for the analyses presented in this paper. ECHILD is available for re-use through the Office for National Statistics Secure Research Service (ONS SRS).

## Data availability statement

The ECHILD database is made available for free for approved research based in the UK, via the ONS Secure Research Service. Enquiries to access the ECHILD database can be made by emailing. Researchers will need to be approved and submit a successful application to the ECHILD Data Access Committee and ONS Research Accreditation Panel to access the data, with strict statistical disclosure controls of all outputs of analyses.

---

**Supplementary Table 1.**
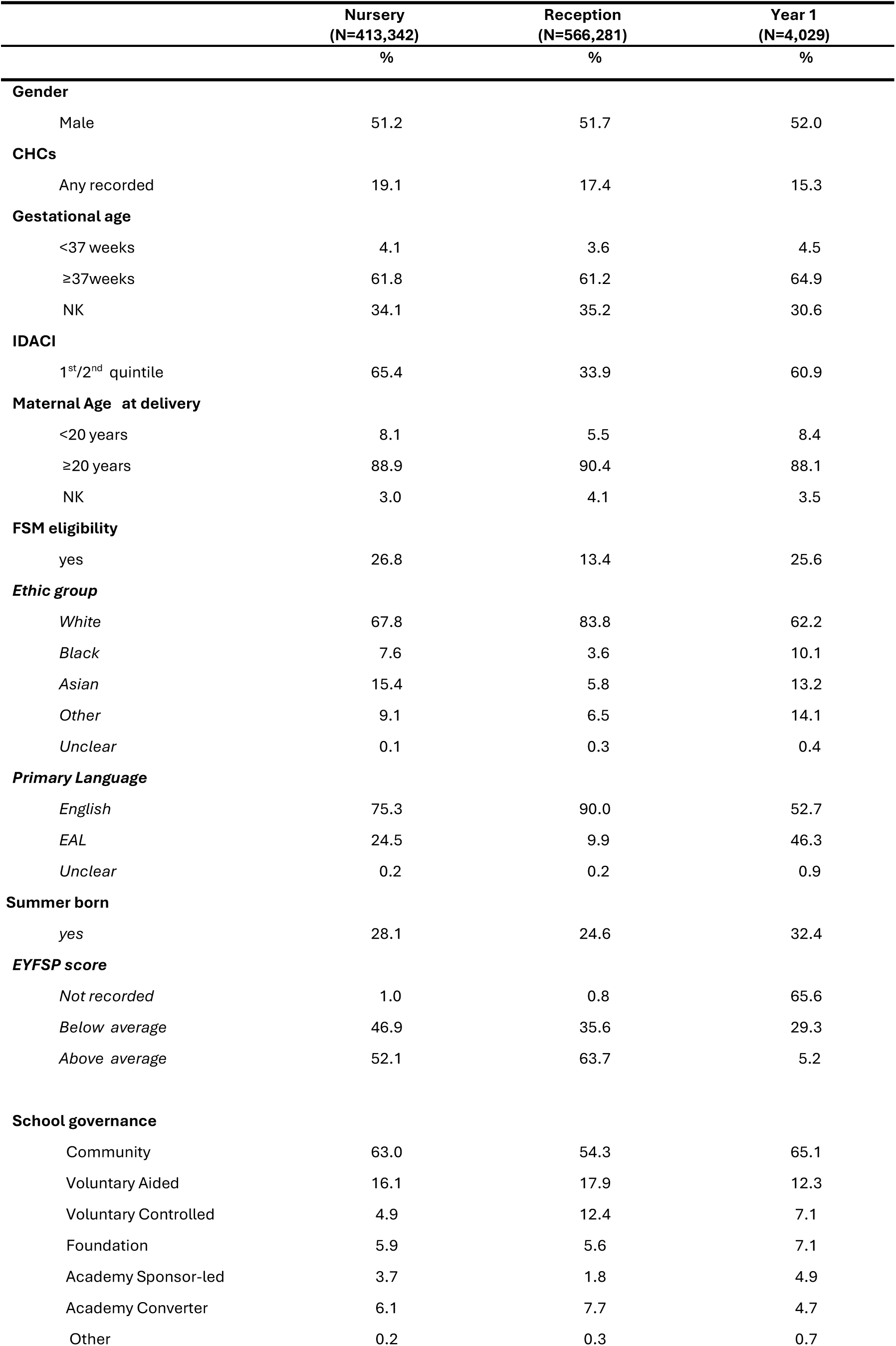
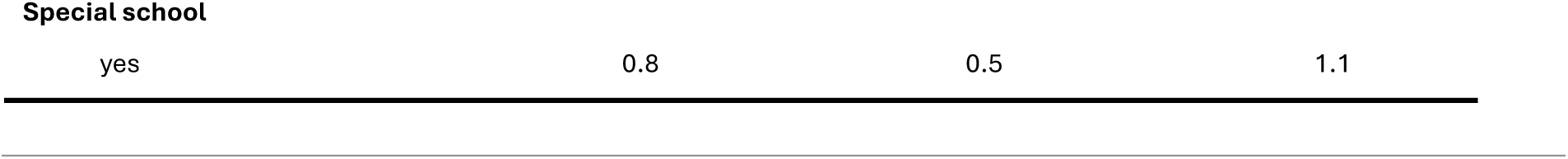
Distribution of children characteristics by stage at entry into state education; HOPE cohort of children born in 2006-08; N=983,652.

**Supplementary Table 2.** Comparison of fitted latent class models, N=983,652.

| **Number of**<br>**classes** | **Log-likelihood** | **AIC** | **BIC** | **Class percentages** | **Comments** |
| --- | --- | --- | --- | --- | --- |
| 1 | -2,623,224 | 5,246,459 | 5,246,530 | 100% | Reference model |
| 2 | -1,528,110 | 3,056,246 | 3,056,399 | 80.3%<br>19.7% | 2 classes better than 1<br>class |
| 3 | ----- | ----- | ----- | ----- | Did not converge |
| 4 | **-1371429** | **2,742,913** | **2,743,231** | **77.1%**<br>**11.8%**<br>**5.7%**<br>**5.6%** | **Best fitting model** |
| 5 | ----- | ----- | ----- | ----- | Did not converge |

**Supplementary Figure 1.**
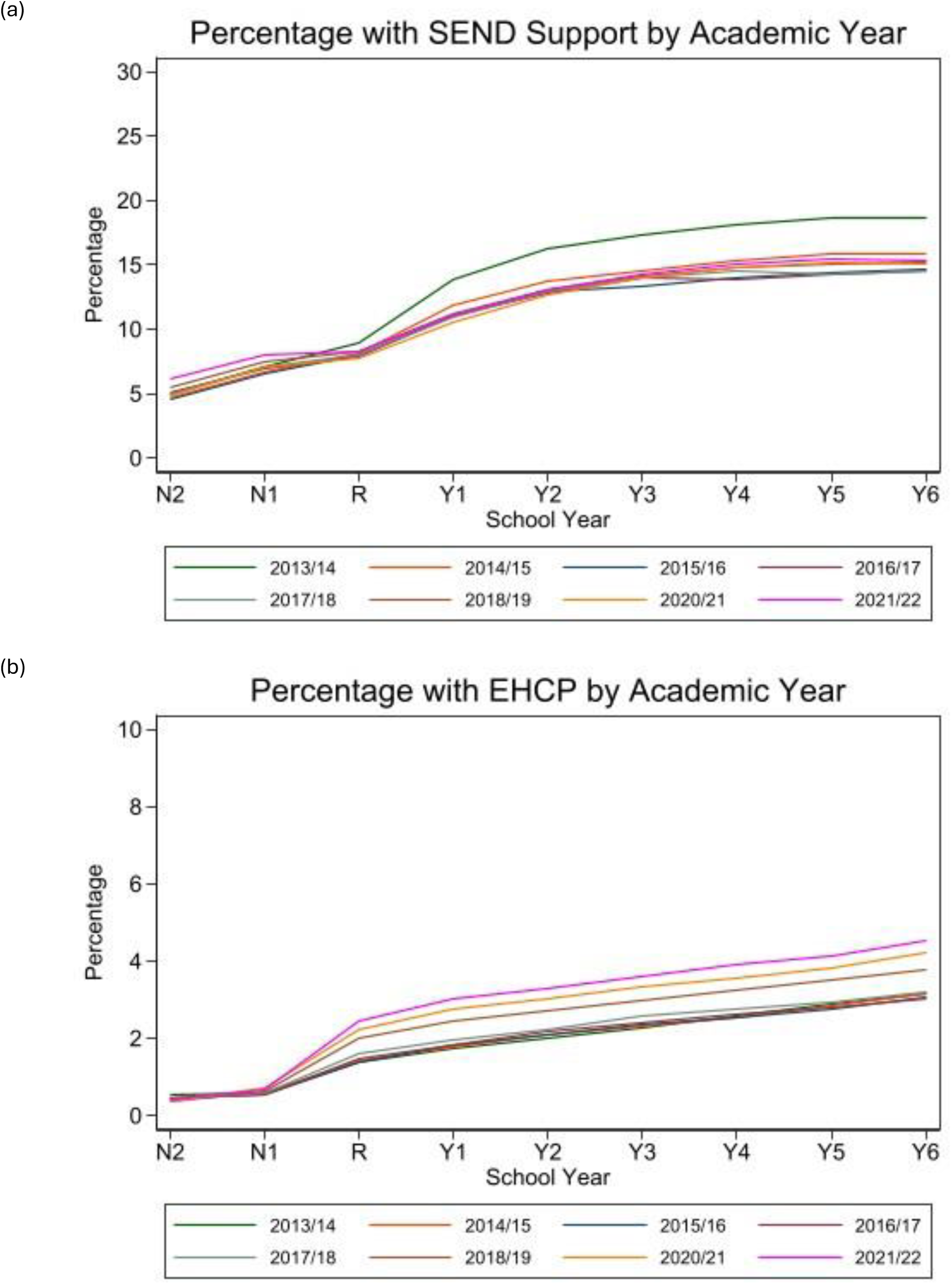
Percentage of SEND support and EHCP from Nursery to Year 6 by academic year; NPD database

**Supplementary Figure 2.**
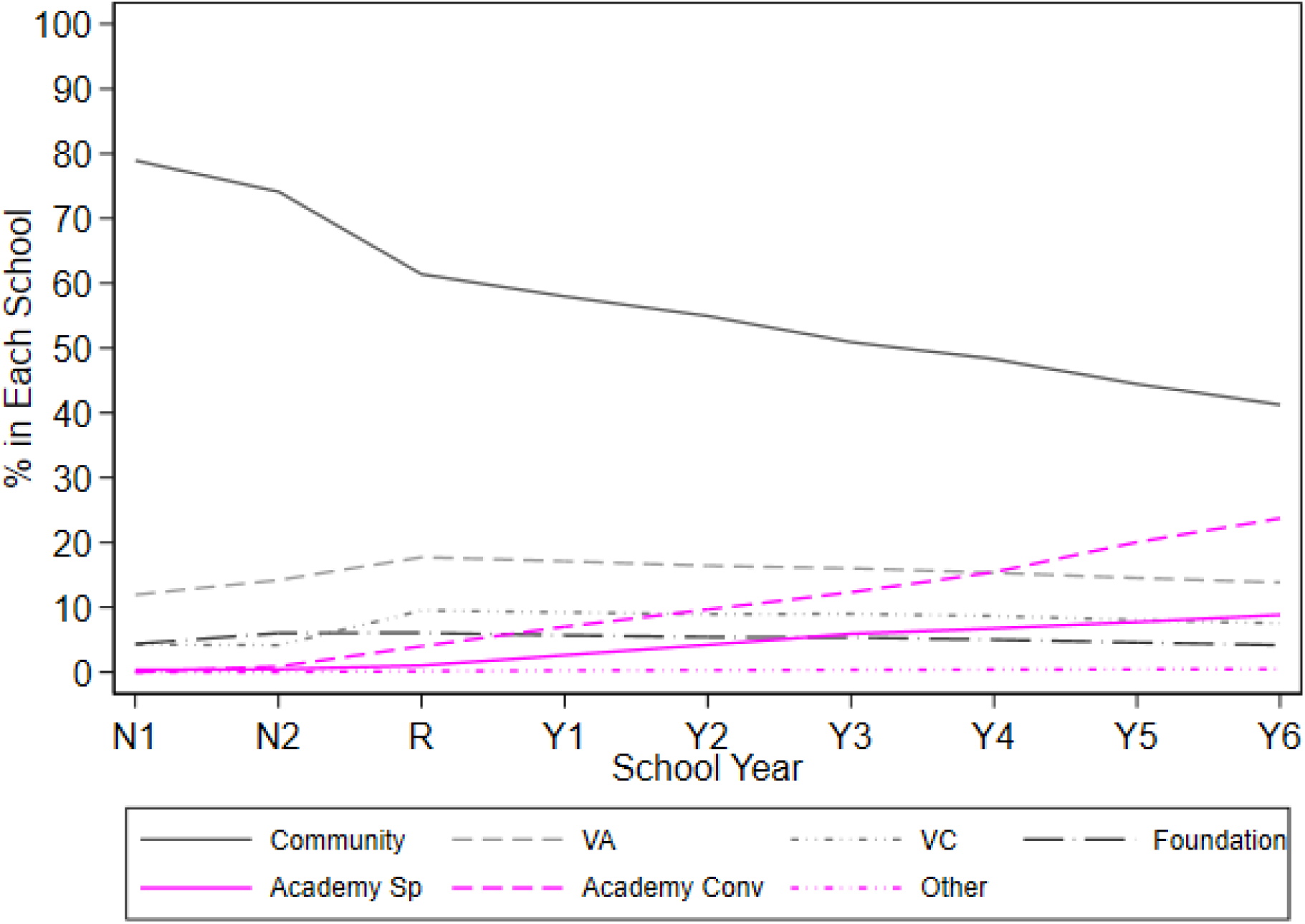
Percentage of children attending different school governance types from Nursery to Year 6 by academic year; HOPE cohort of children born in 2006-08; N=983,652

## Appendix 1: Glossary

Glossary of common terms used in the paper (for more details of terms used in the HOPE study see Gilbert et al, 2025).

| School governance and types of primary schools |  |
| --- | --- |
| **Local Authority Maintained Schools**<br>Local authority (see below) maintained schools are fully owned and operated by local authorities. They follow the national curriculum, are closely connected to their local communities, and may offer additional services such as childcare. Maintained schools can be named in a Statement of SEN or an EHCP, following consultation with the local authority, and must comply with government regulations on admissions, exclusions, and SEND provision.[10] |  |
| **Community school** | A local authority-maintained, secular school. The local authority manages admissions, staffing, and owns the buildings. |
| **Voluntary Aided school** | These schools are typically faith-based, though admissions are open to all applicants. They are jointly funded by the local authority and a supporting religious body, such as the Roman Catholic Church. The governing body employs staff, sets admissions policies, and contributes to building and maintenance costs. School premises are usually owned by a charitable foundation. |
| **Voluntary Controlled school** | Similar to voluntary aided schools, these are mostly faith-based but are fully funded by the local authority. While the local authority is responsible for admissions and employs school staff, it consults with the supporting body when setting the admissions policy. The land and buildings are typically owned by a charitable foundation. |
| **Foundation school** | Funded by the local authority but are managed by their governing body, which acts as the admission authority. The governing body is responsible for employing staff and overseeing admissions. The land and buildings are owned either by the governing body or a charitable foundation. |
| **Academy (Sponsor-led)** | State-funded school set up to replace underperforming schools. Overseen by an external sponsor (e.g., trust, charity). Operates independently from local authorities with freedom over curriculum, staffing, and budget. |
| **Academy (Converter)** | A higher-performing school that voluntarily converts to academy status. Funded directly by the Department for Education and has autonomy over operations and curriculum. |
| **Special school** | Special primary schools in England serve pupils with a broad spectrum of special educational needs and disabilities (SEND; see section 5 in this glossary), including those with severe and complex disabilities. These schools provide tailored support and specialised learning environments designed to meet the needs of children who require more intensive or individualised interventions than those typically available in mainstream educational settings.[7] Almost all children in a special school have an education, health and care plan (EHCP).[8] Special schools may be maintained by local authorities or operate as academies, including both sponsor-led and converter academies. They may be either faith-based or secular, and in exceptional cases, may operate as independent (private) schools.[9] |
| **Types of SEND Provision** |  |
| **SEN Support** | A less intensive form of SEND provision received by most pupils identified as having special educational needs or disabilities. It is school-led and school-funded and may include short-term interventions such as speech and language therapy, or additional support with reading and learning.[5] |
| **Education, Health and Care Plan (EHCP)** | A more intensive form of SEND provision for children and young people whose needs require legally enforceable support. EHCPs are assessed and partly delivered by the local authority and can remain in place until the age of 25.[5] They can be delivered in both mainstream and special schools, depending on the individual's needs. Funding is primarily provided by the local authority where the child or young person resides, with additional contributions from the educational setting they attend (which may be located in a different local authority area). EHCPs replaced Statements of SEN following the 2014 education reforms.[37] |
| **Other terms** |  |
| **Local Authority** | LA was defined by the middle layer super output area 2001 or 2011 of each child's residential address reported in the pupil-level school censuses (NPD) at entry into Year 1 and mapped to one of the 150 upper tier LAs in England as defined in 2021 (excluding Isle of Scilly and City of London due to small pupil size). LAs are local government organisations that are responsible for services in a defined area and its resident population, such as education, transport and social care.<sup>15</sup> LAs are responsible for allocating funding for SEND s |

